# Pregnancy Desire and Pregnancy Attempt: Why Words Matter in Reproductive Research — A Nationwide cross-sectional Cohort Study

**DOI:** 10.64898/2026.03.17.26348589

**Authors:** Rayan Kabirian, Raphaëlle Bas, Agathe Chabassier, Clara Sebbag, Christine Rousset-Jablonski, Angelique Bobrie, Florence Coussy, Marie Préau, Marc Espie, Elise Dumas, Fabien Reyal, Guillemette Jacob, Floriane Jochum, Anne-Sophie Hamy, Seintinelles research network

## Abstract

**Objective:** To quantify the gap between pregnancy desire and pregnancy attempts among young women with and without a history of breast cancer (BC), and to identify factors associated with this gap.

**Design:** Cross-sectional cohort study.

**Setting:** The FEERIC study, conducted in France.

**Population:** Women aged 18–43 years without or with prior BC filling inclusion forms of a collaborative study.

**Methods:** Pregnancy desire was assessed by self-report (“Do you currently desire a pregnancy?”). Attempt was defined as engaging in unprotected intercourse with the intention to conceive. The pregnancy desire–attempt gap was defined as expressing a desire for pregnancy without actively trying to conceive. Logistic regression was used to evaluate associated demographic, clinical, and treatment-related factors.

**Main outcome measures:** Prevalence of the pregnancy desire–attempt gap and predictors of this gap among BC survivors.

**Results:** Of 4,351 participants (517 with BC and 3,834 controls), 735 (16.9%) reported a pregnancy desire with 54% attempting conception and 46% who did not. The desire–attempt gap was significantly more frequent in women with a history of BC (OR=1.62, 95%CI[1.15–2.30]). Among BC survivors, younger age (<30years), nulliparity, being single, and ongoing endocrine therapy were independently associated with the gap, whereas prior chemotherapy or trastuzumab were not.

**Conclusions:** Nearly half of women declaring desiring pregnancy do not initiate pregnancy attempts, with a larger gap among BC survivors. These findings highlight the need to explore both medical barriers and psychosocial determinants underlying this gap and underscore the importance of refining the language used in reproductive research.

**Funding:** This study was supported by “SHS INCa” grant no.2016-124 and is part of a research project on young women funded by Monoprix*.

## Introduction

Many women, at some point in their lives, express a desire to have children, but this desire does not always translate into a concrete attempt to conceive (1). “Desire” reflects an intrinsic aspiration, whereas an “attempt” or “intention” implies a concrete, measurable plan. These terms are often used interchangeably in publications, frequently leading to confusion as this terminological imprecision can obscure a substantial gap between what women want and what they actually do (2).

Breast cancer (BC) is the most common cancer in women of reproductive age. Advances in treatment and survival have shifted attention toward long-term survivorship issues, including fertility and pregnancy after BC (3,4). Concerns about the risk of recurrence have been alleviated by large studies showing pregnancy to be safe, regardless of hormone receptor, *HER2*, or BRCA mutation status (5–7). However, several medical (treatment-related delays, tamoxifen contraindications), psychological (fear of recurrence, relationship difficulties) (8–10), and biological (age-related fertility decline, chemotherapy-induced ovary damage) factors (11,12) may modify fertility and pregnancy plans.

For women with a history of BC, the distinction between a desire for pregnancy and a real intention to conceive is particularly important. There is evidence to suggest that women with a history of BC have a significantly lower likelihood of pregnancy than the general population(13), but it remains unclear whether these low pregnancy rates are due to a lack of desire for pregnancy, a low frequency of attempts to conceive or genuine infertility.

The FEERIC study was designed to compare fertility, pregnancy, and contraception outcomes in young women who have survived BC and matched women with no history of cancer. Previous analyses showed that the overall prevalence of contraceptive use after three years of follow-up was similar in women with a history of BC and in controls and that time-to-pregnancy did not differ significantly from that in controls in the women actively seeking to conceive who became pregnant.

Here, we specifically examine the relationship between the desire for pregnancy and attempts to conceive, with the aim of identifying factors associated the gap between these two entities.

## Methods

### Study design and setting

The FEERIC (Fertility, Pregnancy, Contraception After Breast Cancer in France) study is a nationwide, prospective, exposed–unexposed cohort study conducted through the Seintinelles collaborative research platform. Seintinelles is a social network created in 2012 to facilitate the recruitment of participants for studies on cancer by connecting researchers with volunteers across France. Recruitment for FEERIC took place between March 13, 2018, and June 27, 2019. Participants completed baseline and follow-up questionnaires, we focused for this particular topic on baseline ones.

The study protocol was approved by the Seintinelles scientific board (December 7, 2015) and the Sud Ouest Outre Mer II ethics committee (October 5, 2017, reference 2017:A02181-52). All participants provided electronic informed consent. The design of this study has been described in detail elsewhere (14–16).

### Study population

Women aged 18–43 years were eligible for participation in this study. The exposed group comprised women with a history of localized, relapse-free BC (invasive or *in situ*) who had completed primary treatment (surgery, chemotherapy, radiotherapy) at inclusion. Women receiving adjuvant endocrine therapy or trastuzumab at enrollment were eligible. The unexposed group comprised women of the same age range, free of breast or other cancers. The exclusion criteria for both groups were prior hysterectomy, bilateral oophorectomy, or bilateral salpingectomy. Women were recruited via the Seintinelles platform and through eight French cancer and oncofertility centers (Institut Curie [Paris and Saint-Cloud], Hôpital Saint-Louis [Paris], Hôpital Cochin [Paris], Centre Léon Bérard [Lyon], Centre Oscar Lambret [Lille], Centre Hospitalier Régional Universitaire [Lille], and Institut Bergonié [Bordeaux]).

### Data collection

Data were collected through self-administered online questionnaire hosted on the Seintinelles platform. At baseline, participants reported sociodemographic characteristics, medical and reproductive history. We considered two specific variables: pregnancy desire and pregnancy attempts. **Pregnancy desire** was assessed with the question: “Do you currently desire a pregnancy?” with the options: “yes”, “no”, or “don’t know”. Women who answered “yes” were considered to desire a pregnancy. **Pregnancy attempts** were defined as engaging in unprotected sexual intercourse with the intention to conceive, as assessed with the question: “Are you currently engaging in unprotected intercourse with the intention of becoming pregnant?”, with the options: “yes”, “no”, or “prefer not to answer”. Women who answered “yes” were considered to be actively attempting to fall pregnant.

### Outcomes

The primary outcome was the pregnancy desire-intention gap, the mismatch between reproductive desires and behavior at study inclusion, defined as the proportion of women reporting a desire for pregnancy who were not actively attempting to conceive. Secondary objectives included the identification of demographic, clinical, and treatment-related factors associated with this gap.

### Statistical analysis

Descriptive statistics are reported as frequencies and percentages for categorical variables, and medians with standard deviations (SD) for continuous variables. Comparisons between groups were performed with Chi-squared or Fisher’s exact tests for categorical variables, and Student’s *t*-tests or Wilcoxon–Mann–Whitney tests for continuous variables, as appropriate.

We first assessed factors associated with the pregnancy desire-intention gap in univariable analyses. The variables considered included sociodemographic (age, marital status, socioprofessional category, educational level), lifestyle (BMI, smoking), reproductive (parity, menstrual cycles), and clinical (prior chemotherapy, trastuzumab, endocrine therapy, hereditary predisposition, comorbid conditions) variables. Variables with a p-value for association <0.1 in univariable analysis were included in multivariable analysis. Multivariable logistic regression models were then fitted separately for the total study population and in the subgroup of women with a history of BC to identify independent predictors of the pregnancy desire-intention gap.

All analyses were performed with R software (R Foundation for Statistical Computing, Vienna, Austria). A *p-*value <0.05 in two-tailed tests was considered statistically significant.

## Results

### Study population

In total, 4,351 women were included in the FEERIC study, 517 (11.9%) of whom had been treated for BC, the other 3,834 (88.1%) being controls with no history of cancer. Mean age at inclusion was 33.5 years (SD: 4.6 years). The characteristics of the women at baseline are summarized in **Table 1**. Most participants had been educated to university level (3804/4351, 87.4%), were non-smokers (3618/4351, 83.1%), and reported no comorbid conditions (3014/4351, 69.3%). About half already had children (49.4%), and three-quarters were in a relationship (75.9%). Among women with a history of BC, median time since diagnosis was 34.9 months, 424/517 (82.0%) had received chemotherapy, 256/517 (49.5%) have ongoing endocrine therapy, and 126/517 (24.4%) were on trastuzumab.

**Table 1.**
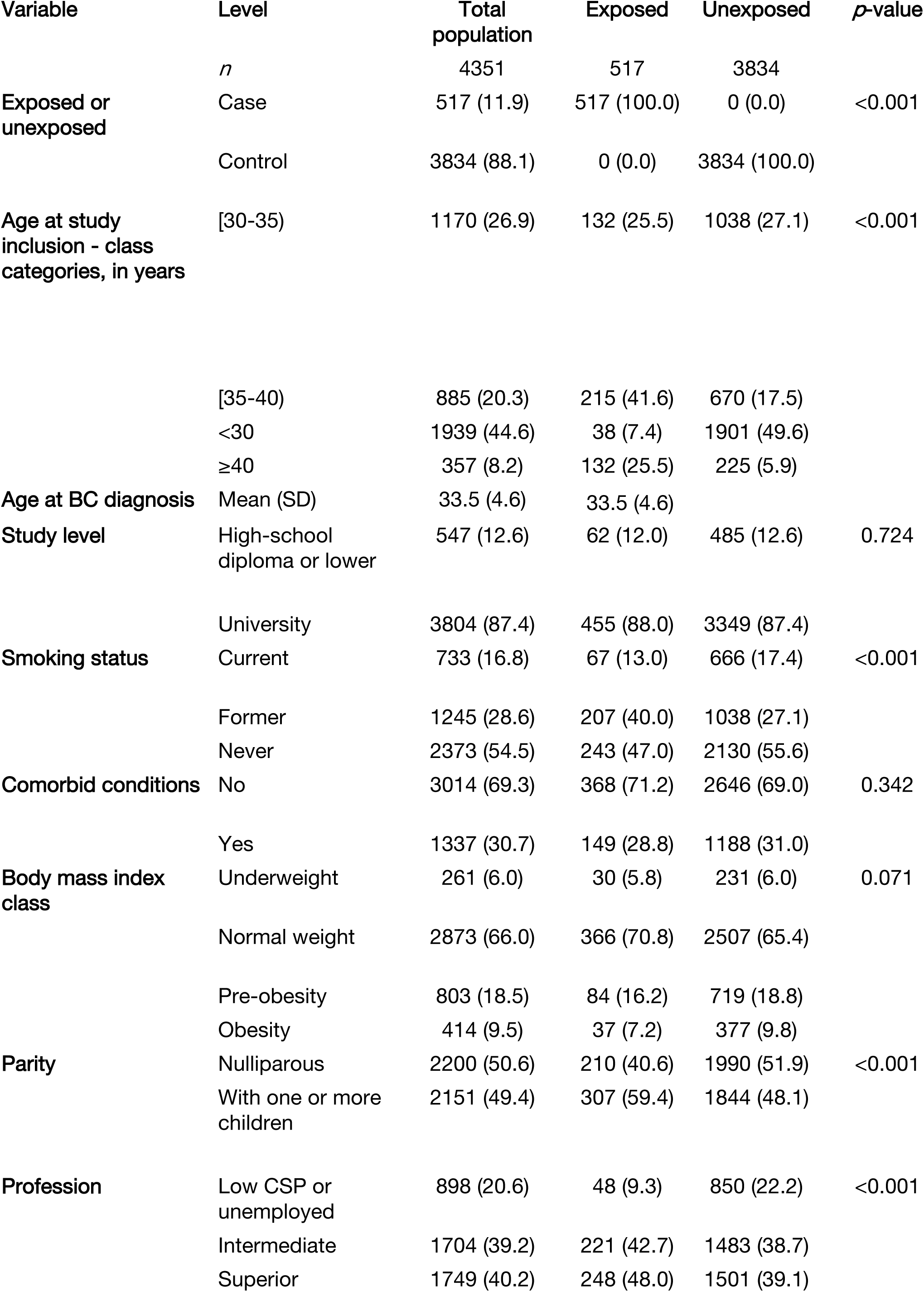
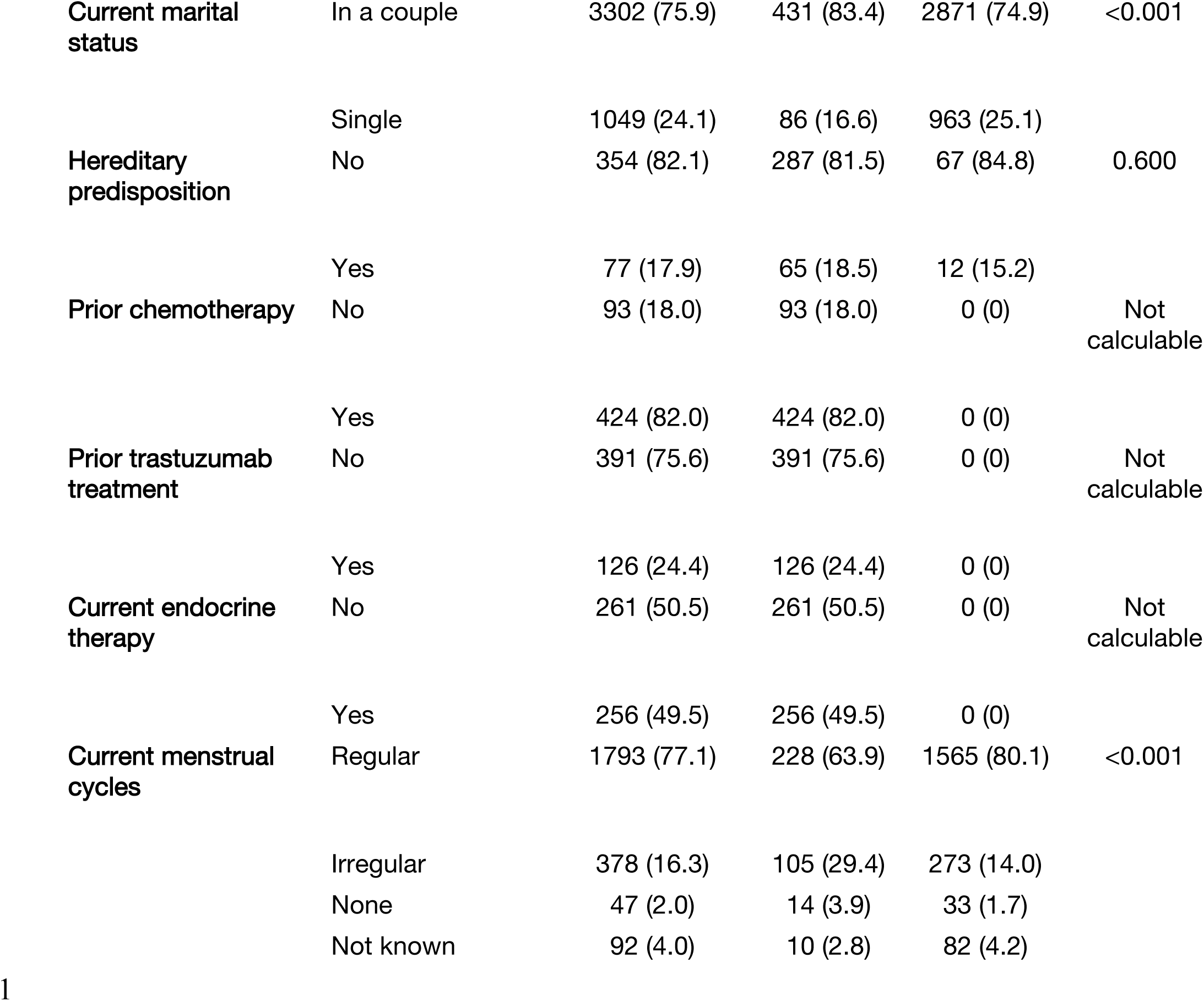
Baseline characteristics of the study population (*N*=4,351). Values are presented as *n* (%)

### Pregnancy desire

In response to the question about whether they desired a pregnancy, 3,239 women (74.4%) reported no desire for pregnancy, 735 (17.0%) reported a desire, and 377 (8.7%) were uncertain. (**Figure 1**).

**Figure 1.**
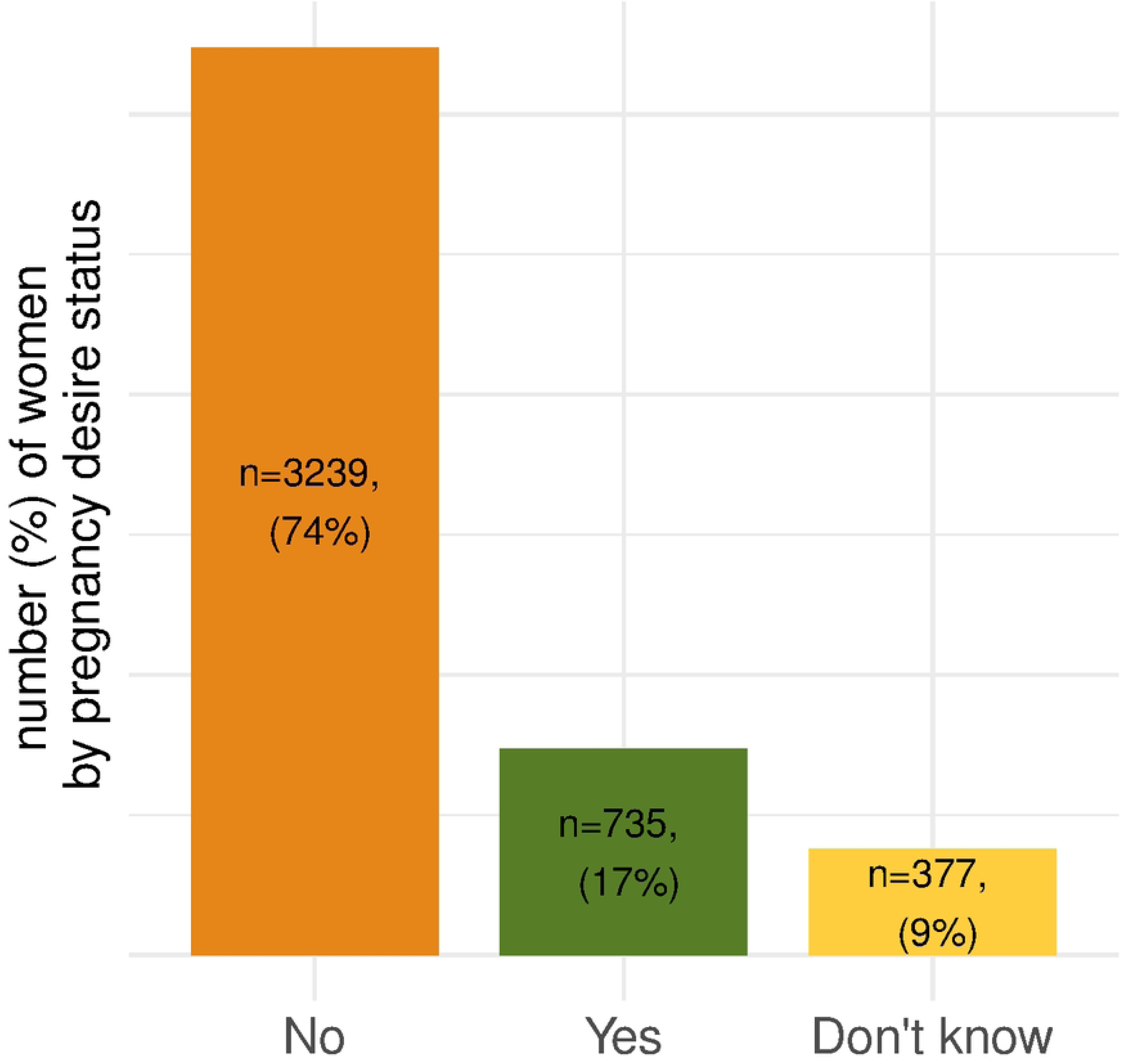
Distribution of pregnancy desire at inclusion among women in the FEERIC study (*N*=4,351).

### Pregnancy attempts

None of the women who declared having no desire for a pregnancy or being uncertain about their desire reported attempting to conceive. In total, 397 (54.0%) of the 735 women in the cohort who expressed a desire to become pregnant reported actively attempting to conceive, whereas the other 338 (46.0%) women expressing a desire for pregnancy did not report attempting to conceive. In the group of women with a history of BC, 178 women expressed a desire to become pregnant, only 66 (37.1%) reported actively attempting to conceive, the 112 (62.9%) reporting no active attempt to conceive (**Figure 2**).

**Figure 2.**
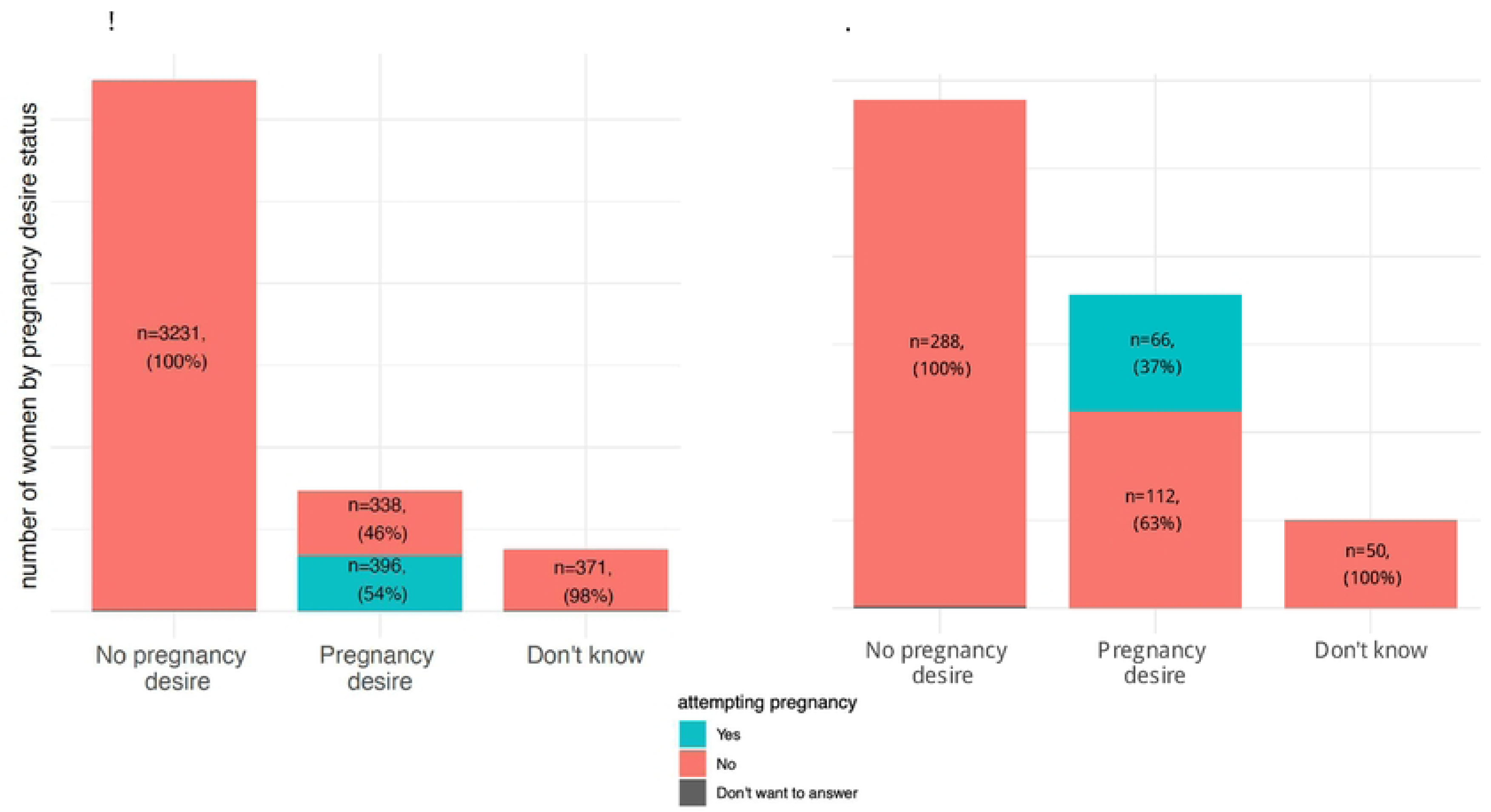
Distribution of women attempting to conceive at inclusion according to stated pregnancy desire status in the FEERIC study (*N*=4,351) and in women with a history of BC (*N*=517). **B. Total cohort.** In total, 15 patients preferred not to say whether or not they were attempting to conceive. These patients are shown in gray, with the following distribution concerning their responses relating to the desire for a pregnancy: Don’t know *n*=6, 2%; No *n*= 8, 0%, Yes *n*= 1; 0%). **C. Women with a history of breast cancer.** One patient who expressed an absence of desire for pregnancy did not wish to answer the question about pregnancy attempts

### Factors associated with the pregnancy desire-intention gap

In the 735 women with a desire to become pregnant, several factors were associated with a pregnancy desire-intention gap in univariable analyses (**Table 2 and Figure 3**). Such a gap was more frequently observed in women with a history of BC (odds ratio OR=1.62, 95% CI: 1.15–2.30), younger women (vs. ≥40 years (reference class), [35-40), OR=2.18, 95% CI: 1.07–4.69), [30-35), OR=3.00, 95% CI: 1.51–6.31), and <30, OR=4.84, 95% CI: 2.45–10.19), nulliparous women (OR=1.41, 95% CI: 1.05–1.91), women from lower socioprofessional categories (vs. superior, OR=1.20, 95% CI: 0.76–1.93), single women (OR=4.14, 95% CI: 2.47–7.29), and women on ongoing endocrine therapy (OR=10.03, 95% CI: 4.59–24.53). Conversely, education level, comorbidity, hereditary predisposition, and prior trastuzumab treatment or chemotherapy were not associated with a pregnancy desire-intention gap.

**Figure 3.**
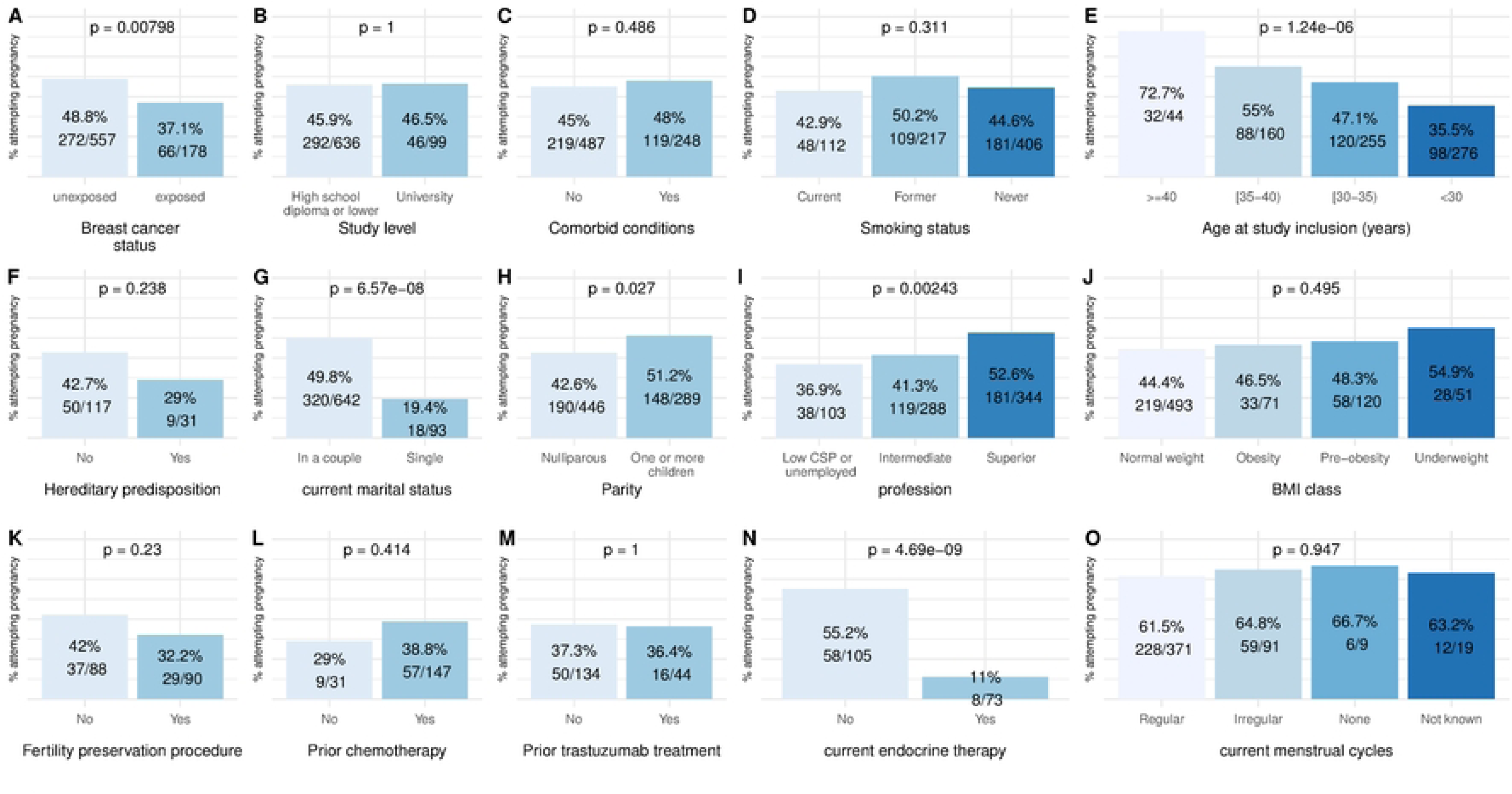
Association between the characteristics of the women and discordance between pregnancy desire and pregnancy attempts in women reporting a desire for pregnancy (*n*=735). Each panel (A–N) displays the proportion of women attempting pregnancy among those declaring a desire for pregnancy, according to: (A) breast cancer status, (B) education level, (C) comorbidy, (D) smoking status, (E) age at study inclusion, (F) hereditary predisposition, (G) current marital status, (H) parity, (I) profession, (J) BMI class, (K) fertility preservation procedures, (L) prior chemotherapy, (M) prior trastuzumab treatment, (N) current endocrine therapy, and (O) current menstrual cycles. *P*-values from chi-square tests are indicated.

**Table 2.**
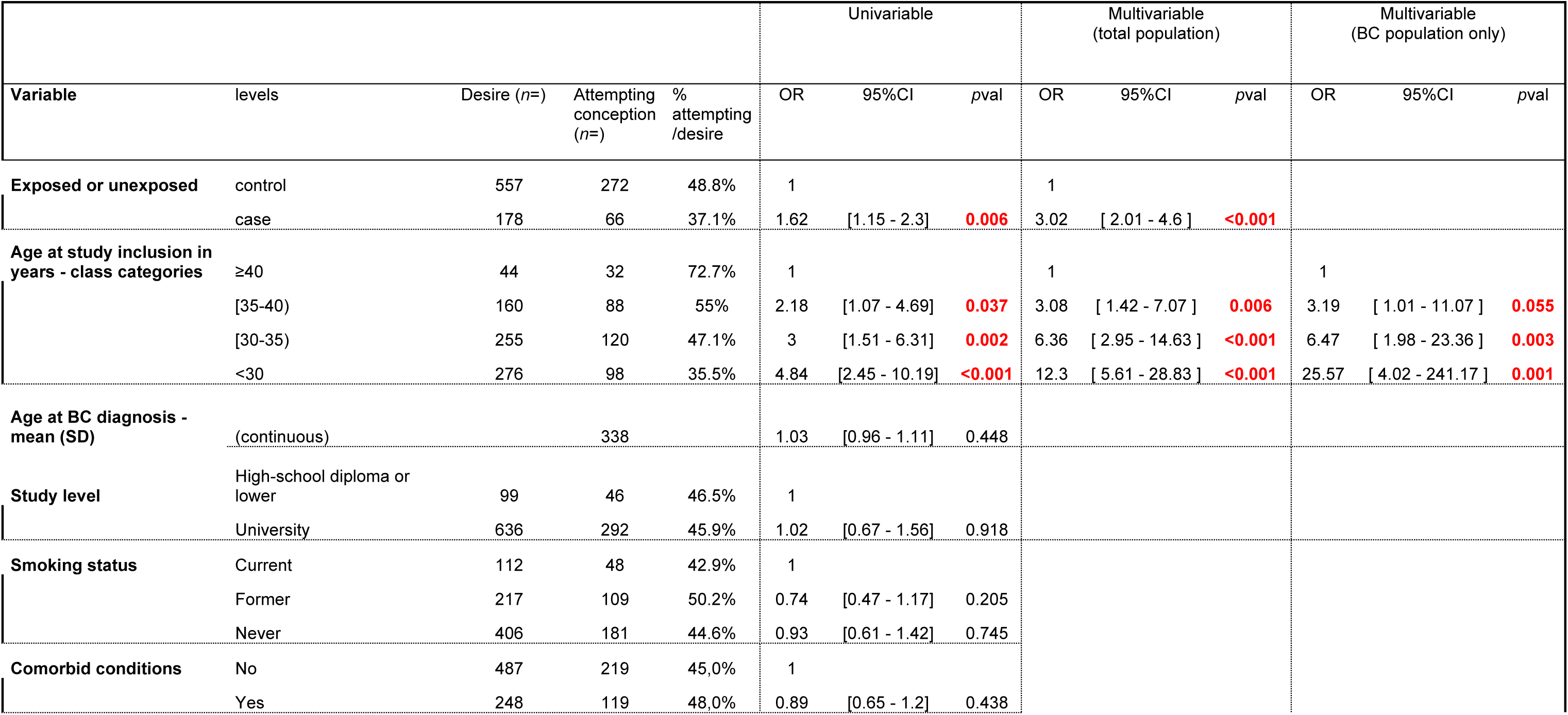
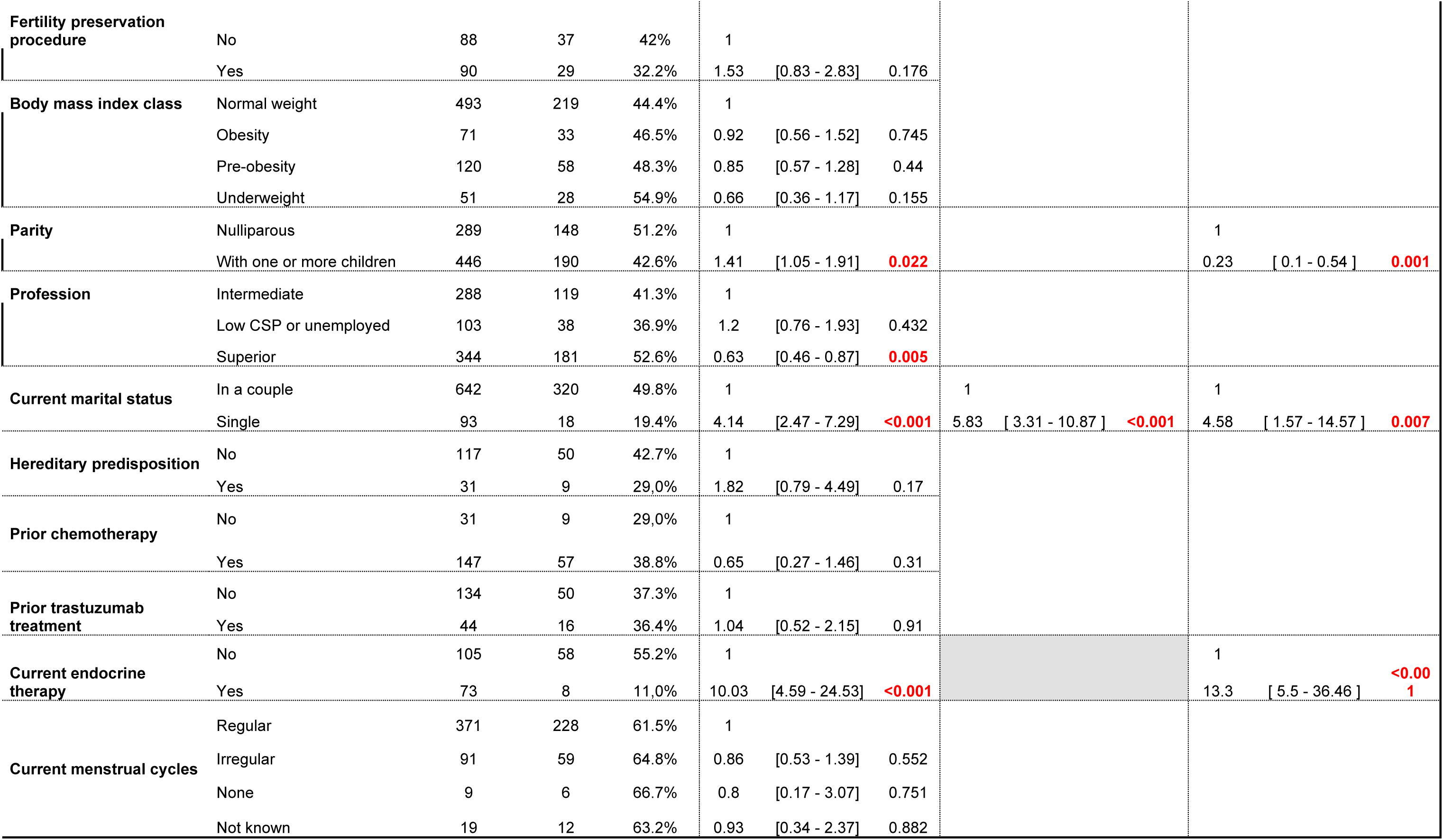
Factors associated with the pregnancy desire-intention gap — discord between the desire for a pregnancy and active attempts to conceive. *Results are presented as crude and adjusted odds ratios (OR) with 95% confidence intervals (CI) from univariable and multivariable logistic regression models for the total cohort (N=4,351) and for the cohort of women with a history of BC (N=517)*

Three factors remained independently associated with the pregnancy desire-intention gap after multivariable analysis adjusted on all variables with p<0.1 in univariate analysis: prior BC (adjusted OR (aOR)=3.02, 95% CI/ 2.01–4.60, *p*<0.001), younger age at inclusion (vs ≥40 years: aOR=3.08, 95% CI: 1.42–7.07 for [35-40); aOR=6.36, 95% CI: 2.95–14.63 for [30-35), and aOR=12.3, 95% CI 5.61–28.83 for <30), and being single (aOR=5.83, 95% CI: 3.31–10.87, *p*<0.001).

Multivariable analysis on the specific subgroup of women with a history of BC (*n*=517) identified age at inclusion (vs ≥40 years: aOR=3.19, 95% CI: 1.01–11.07 for [35-40), aOR=6.47, 95% CI: 1.98–23.36 for [30-35), and aOR=25.57, 95% CI: 4.02–241.2 for <30), parity (aOR=0,23, 95%CI: 0.1-0.54 for parous vs. nulliparous), being single (aOR=4.58, 95% CI: 1.57–14.57, *p*=0.007), and current endocrine therapy (aOR=13.3, 95% CI: 5.5–36.5, *p*<0.001) as factors significantly associated with the pregnancy desire-intention gap (**Table 2**).

## Discussion

In this large cross-sectional cohort of women from the FEERIC study, we observed that only about half of those reporting a for pregnancy were actively attempting to conceive, highlighting the existence of a substantial pregnancy desire–intention gap. Our findings are consistent with the small number of studies on this subject to date, which have reported that a desire for pregnancy does not always translate into action (2).

In the total study population, being younger, single, and having a history of BC were the strongest predictors of this gap. The influence of age is consistent with previous demographic studies reporting that younger women often defer attempts to conceive despite reporting a desire for pregnancy, reflecting educational, professional, or relationship priorities (17–19). Being single emerged as one of the strongest predictors, underscoring the role of relational context in shaping the translation of a desire for pregnancy into attempts to conceive, consistent with previous qualitative studies highlighting how partner availability and relationship dynamics influence reproductive decision-making (20,21)

We found that exposure to BC was a strong risk factor for pregnancy desire-attempt gap, with a risk of such a gap in this group of women almost three times higher than that in cancer-free women. A history of cancer often reshapes reproductive trajectories, with medical concerns and social factors delaying or preventing attempts to conceive.

Younger women, those without children, and those on endocrine therapy were particularly likely to display a pregnancy desire-intention gap, suggesting that both medical barriers such as appropriate clinical guidance constraints, including the contraindication of tamoxifen during pregnancy, and psychosocial determinants were at work. Interestingly, classical medical factors, such as prior chemotherapy, trastuzumab treatment and comorbid conditions, were not independently associated with pregnancy desire-intention gap, suggesting that survivorship and ongoing treatment (especially endocrine therapy) have a greater influence than past exposures.

One of the key contributions of this study lies in its underscoring of the importance of terminology and measurement. The interchangeable use of “desire,” “intention,” and “attempt” in publications in this field has long generated confusion. Pregnancy desire captures an intrinsic aspiration, whereas intention and attempt reflect increasingly concrete steps toward action. In our study, women were explicitly asked about both their desire for pregnancy and their active attempts to become pregnant, making it possible to disentangle these two concepts. This distinction is crucial: the collapsing of these two entities into a single unit is likely to obscure the very gap that demographers and clinicians seek to understand. Previous studies have revealed that many retrospective surveys actually measure the desire for pregnancy rather than the intention to conceive (2) and our findings confirm the need for more precise wording in questionnaire design. Future fertility research should systematically differentiate between these constructs to capture the spectrum of reproductive goals and behaviors more effectively.

In conclusion, this study shows that reproductive aspirations and actions are not always consistent and that both medical and non-medical determinants underlie this mismatch. Beyond clinical risk factors, relationship context and psychosocial conditions determine whether women translate a desire for pregnancy into attempts to conceive, and the ways in which questions are worded can affect the answers given to questions relating to these concepts. Recognizing and measuring these distinctions more precisely represents not only a methodological refinement but also a shift in clinical approach, supporting longitudinal fertility counseling, individualized discussions on reproductive timing (including temporary endocrine therapy interruption when appropriate), and improved structural supports. Future research should continue refining the language used to capture reproductive goals and examine how health systems can better support women in translating desire into action.

## Data Availability

Data cannot be shared publicly because they contain potentially identifiable and sensitive patient information collected within the FEERIC cohort. Access to the data is subject to ethical and regulatory restrictions in accordance with French data protection regulations. Data are available from the FEERIC study coordinating team for researchers who meet the criteria for access to confidential data. Requests for data access can be directed to the FEERIC study investigators via and will be reviewed by the study’s data access committee in accordance with institutional and ethical guidelines.

## Acknowledgments

This study was supported by “SHS INCa” grant no. 2016-124 and is part of a research project on young women funded by Monoprix*.

